# Development and validation of a nomogram for predicting overall survival of patients with ischemic cardiomyopathy and heart failure: a post-hoc analysis of the Surgical Treatments for Ischemic Heart Failure (STICH) trail

**DOI:** 10.1101/2023.10.04.23296574

**Authors:** Pengju Guo, Chang He, Junlei Li, Youxu Jiang, Feng Wang, Jiaxiang Wang, Bin Lin, Deguang Feng

## Abstract

**Purpose:** To establish a nomogram for predicting the overall survival (OS) of patients with ischemic cardiomyopathy and heart failure based on the Surgical Treatment for Ischemic Heart Failure (STICH) trail.

**Methods:** Patients who had valid key variables in the hypothesis 1 were included and randomly divided into the training and validation groups (7:3 ratio). Using Cox proportional hazards model, predictors for the OS in training group were identified and integrated to establish a nomogram for predicting 1-year, 3-year, 5-year, and ten-year survival probability. The nomogram performance was evaluated using Harrell’s concordance index (C-index), time-dependent receiver operating characteristic curve, decision curve analysis, and Kaplan-Meier survival analysis.

**Results:** 940 of 1212 patients who had valid key variables were included. Seven predictors, including treatment type, gender, estimated glomerular filtration rate, Charlson co-morbidity index, 6-minute walk, end-systolic volume index and mitral regurgitation class were identified to establish the nomogram. The C-indices of the nomogram were 0.641 (95% CI: 0.627-0.655) and 0.649 (95% CI: 0.627-0.671) for training and validation groups, respectively. The calibration curves revealed consistency between predicted and observed survival. The area under 1-year, 3-year, 5-year, and ten-year OS receiver operating characteristic curves were 0.634, 0.616, 0.630 and 0.638 in the training group, respectively. Decision curve analysis showed effective net benefits of the model in clinical decision-making. Divided by the cutoff values of prognostic indices, low-risk patients showed better OS than those with high risk in training and validation groups (both p < 0.0001).

**Conclusion:** The current nomogram can effectively predict the OS of patients with ischemic cardiomyopathy and heart failure, provide information about multidisciplinary therapeutic that may prolong the survival time, and serve as a perfect tool in conjunction with the STS and EuroSCORE II risk models in clinical decision making.

## 1. Introduction

Multifaceted strategies to address the control of worldwide prevalence of ischemic cardiomyopathy (ICM) and heart failure, which are projected to affect 8 million US population by 2030, have progressively gained advances during last decades.^1^ Coronary artery bypass grafting (CABG) has become the preferred recommendation for patients with ICM and heart failure in contemporary guidelines with decreasing heterogeneous consensus.^2–4^ However, the realistic concern of triple risk of death within the initial 30 days after randomization among patients who underwent CABG plus medical therapy compared with those received medical therapy alone, identified by the Surgical Treatment for Ischemic Heart Failure (STICH) trial, makes the ultimate clinical decision requiring courage to take a crucial step.^5,6^ This trend toward conservation has resulted the overzealous use of percutaneous coronary intervention (PCI) by nearly triple usage higher than CABG at clinician discretion, but without high-quality evidence.^7,8^ Due to the Society of Thoracic Surgeons (STS) and EuroSCORE II risk models were developed to estimate the early postoperative mortality, the long-term survival in this difficult management but life-prolonging entity can be hardly predicted.^9,10^ Similarly, given the extensive coexisting comorbidities among patients with ICM and heart failure, a dilemma surrounding the implement of the SYNTAX (Synergy Between PCI with TAXUS and Cardiac Surgery) score II 2020 is the interobserver variability and absence of clinical variables, which may largely compromise the generalizability of the model.^11,12^

Landmark results of the STICH trail have driven progress in our understanding of this specific cohort.^5,13^ In the setting of contemporary era of personalized medicine, clinical decision should be patient-centered, which can be achieved through the ascertainment and quantification of survival probability with the engagement of clinicians and patients. However, to the best of our knowledge, there is no specific risk model to predict the overall survival (OS) of patients with ICM and heart failure. Accordingly, we constructed a nomogram based on clinical variables derived from the STICH trail to predict the OS of this specific cohort.

## 2. Methods

### 2.1 Patient source

The current study was a post-hoc analysis of the STICH trail (funded by the National Institutes of Health, NCT00023595), and approved by the National Heart, Lung, and Blood Institute and our institutional review board (2022-KY-1409-002). The data was accessed via the Biologic Specimen and Data Repository Information Coordinating Center. The STICH trail and extended follow-up have previously reported in detail.^5,14^ Among 1212 patients with an left ventricular ejection fraction (LVEF) ≤ 35% and coronary artery disease (CAD) amenable to CABG in hypothesis 1, 940 patients, who had valid preoperative information of age, gender, race, New York Heart Association (NYHA) heart function class, medical history, mitral regurgitation (MR) class, previous revascularization (CABG or PCI), distance of 6-minute walk, LVEF, left ventricular end-systolic volume index (ESVI), Duke CAD index, creatinine, and follow-up information of treatment received, days from randomization to CABG, years of follow-up time, and patients status at the last follow-up, were included in the current study. The detail screening process of patients was shown in Figure 1. The eligible cases were randomly stratified into training and validation groups (7:3 ratio). Each patient provided written informed consent and the investigation complied with the principles outlined in the Declaration of Helsinki.

**Figure 1.**
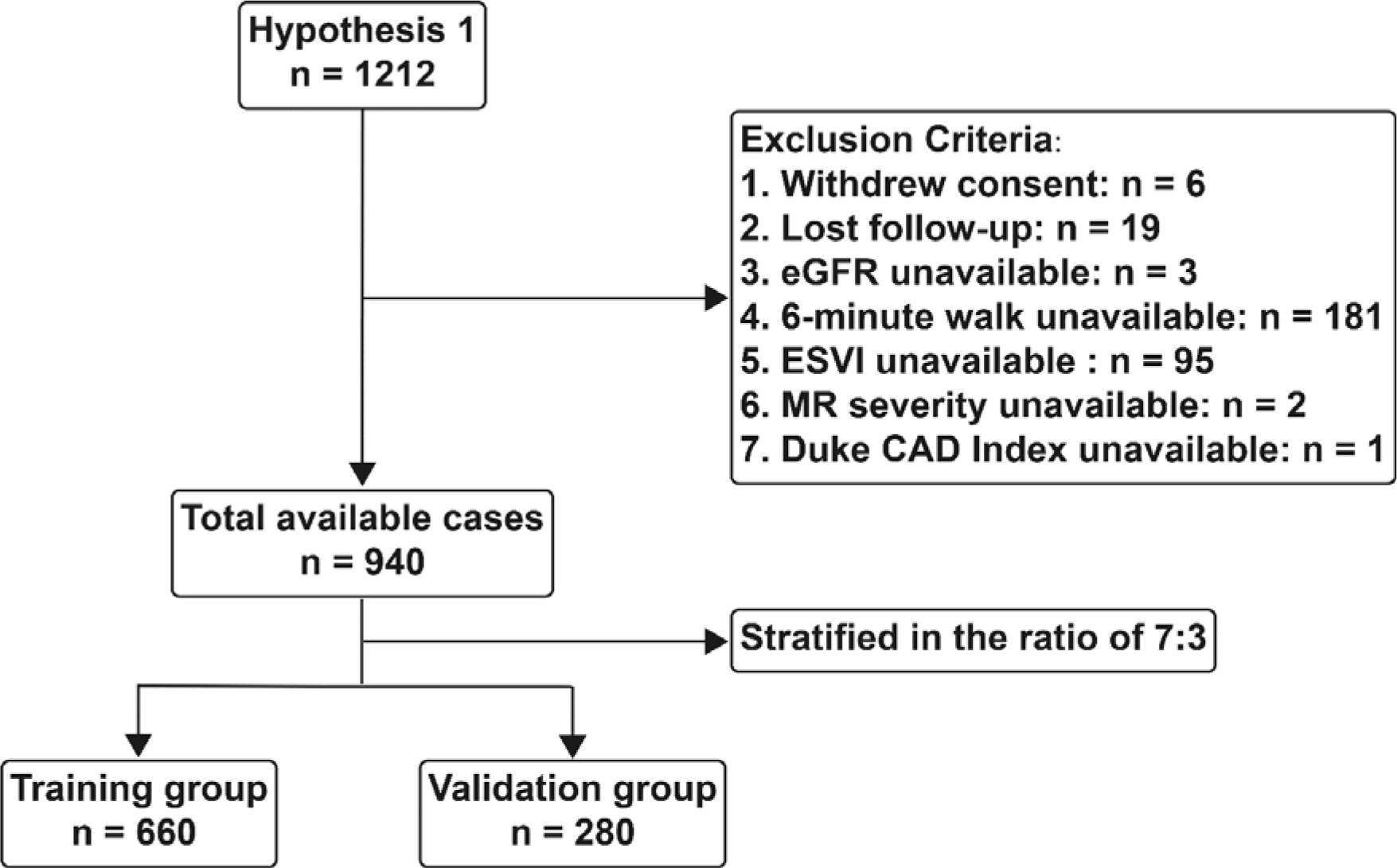
Flow diagram of patient selection. eGFR = estimated glomerular filtration rate; ESVI = end-systolic volume index; MR = mitral regurgitation; CAD = coronary artery disease.

### 2.2 Measurable variables and outcomes

For ease of comparison, age (≤ 50, 51 – 60, 61 – 70, and ≥ 71 years), 6-minute walk (≤ 300 and > 300m), LVEF (≤ 20%, 21 – 25%, 26 – 30%, and 31 – 35%), ESVI (≤ 60, 61 – 90, and ≥ 90 ml/m^2^), and Duke CAD Index (≤ 40, 41 – 70, 71 – 100) were translated into categorical variables. The estimated glomerular filtration rate (eGFR) was calculated using the Chronic Kidney Disease Epidemiology Collaboration (CKD-EPI) equation, and then categorized as > 60 ml/min per 1.73m^2^ or ≤ 60 ml/min per 1.73m^2^.^15^ The Charlson co-morbidity index (CCI) for each patient was accounted based on medical history.^16,17^

Considering the predictive nature of the model, patients who received CABG within 90 days after randomization were assigned to the CABG group (CABG + MED) in the current study, and to medical group (MED) otherwise regardless of whether CABG was implemented exceeding 90 days after randomization. The patient status at the last follow-up was screened and OS was considered as the primary outcome of interest. The duration of survival was defined as the date from randomization to the data of death or last contact.

### 2.3 Statistical analysis

Baseline characteristics were described by frequencies and percentages for categorical variables and by medians (interquartile range [IQR]) for continuous variables. Baseline differences between training and validation groups were compared using Chi-square or Fisher’s exact tests for categorical variable. Univariate and multivariate Cox proportional hazards models were performed to identify the independent predictors for OS in the training group.

Based on the predictors identified by multivariate Cox proportional hazards model, current nomogram was developed to predict the 1-year, 3-year, 5-year, and ten-year survival probability of patients with ICM and heart failure. Harrell’s concordance index (C-index) and time-dependent receiver operating characteristic (ROC) curve were used to evaluated the discrimination of the nomogram for predicting the survival outcomes between different patients. The differences between predicted and actual risks were visualized using calibration curves. Bootstraps with 1000 resamples were performed to calculate the C-indices and plot calibration curves. The net benefit of the nomogram for decision-making was evaluated using Decision Curve Analysis (DCA). The prognostic index of 1-year OS for each patient was calculated using independent predictors and corresponding hazards ratio. Based on the cutoff values of prognostic indices, the training and validation groups were stratified into high-risk and low-risk groups, respectively. Kaplan-Meier survival analysis and log rank test were used to compare the OS between high-risk and low-risk groups.

All statistical analyses were conducted using R software (version 4.2.2, http://www.r-project.org/) and two-sided. Statistical significance was set at P < 0.05.

## 3. Results

### 3.1. Baseline characteristics

A total of 940 patients who had valid key variables were included, and randomly stratified into training group (660 cases) and validation group (280 cases) based on 7:3 ratio. The baseline characteristics of entire cohort, training and validation groups were showed in the Table 1. There was no significant difference between training and validation groups with regard to each variable. The median follow-up time was 7.3 years (IQR: 2.9 - 9.4 years) for training group and 6.7 years (IQR: 3.2 - 9.3 years) for validation group. There were 406 and 177 deaths recorded in the training and validation groups, respectively.

**Table 1.**
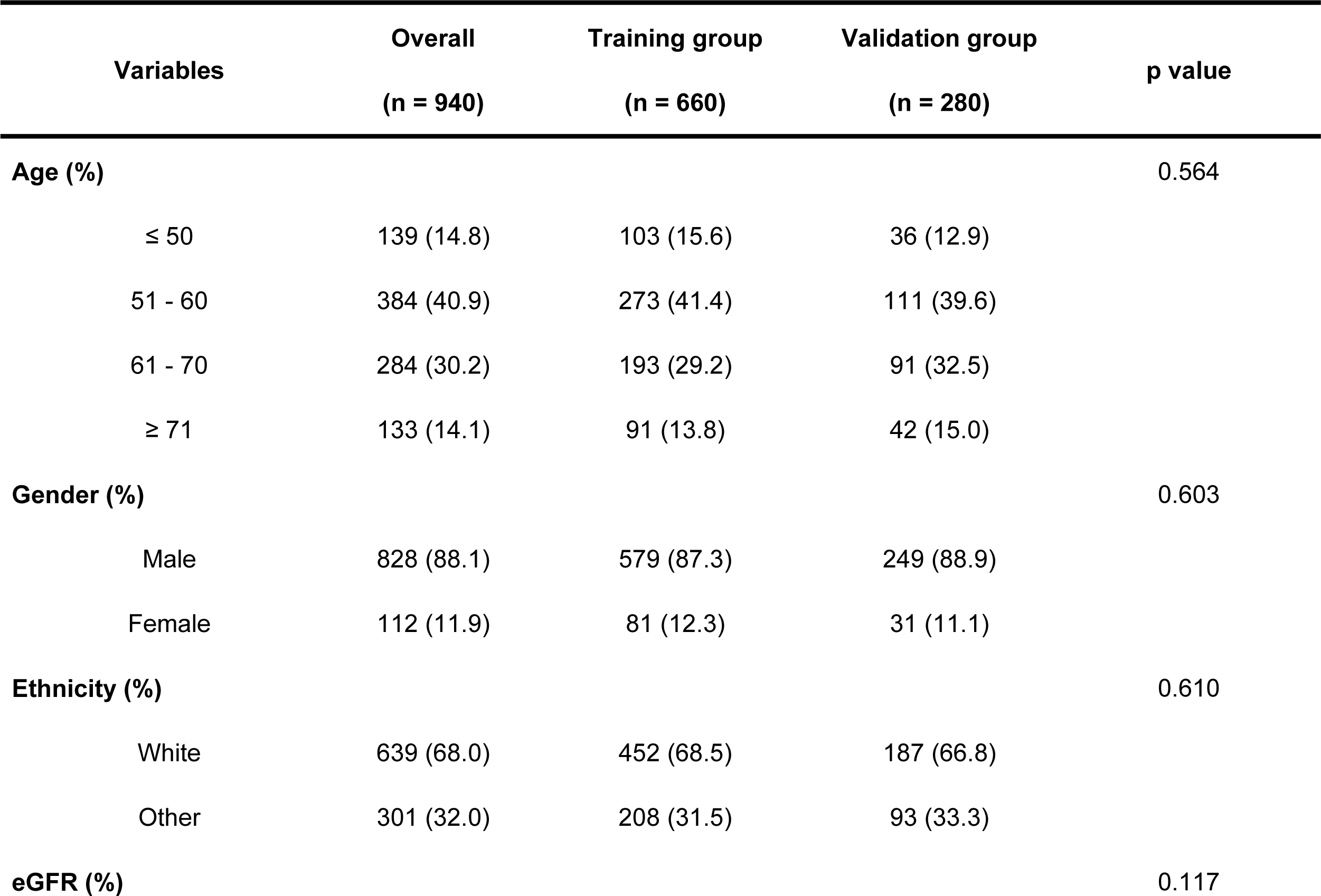
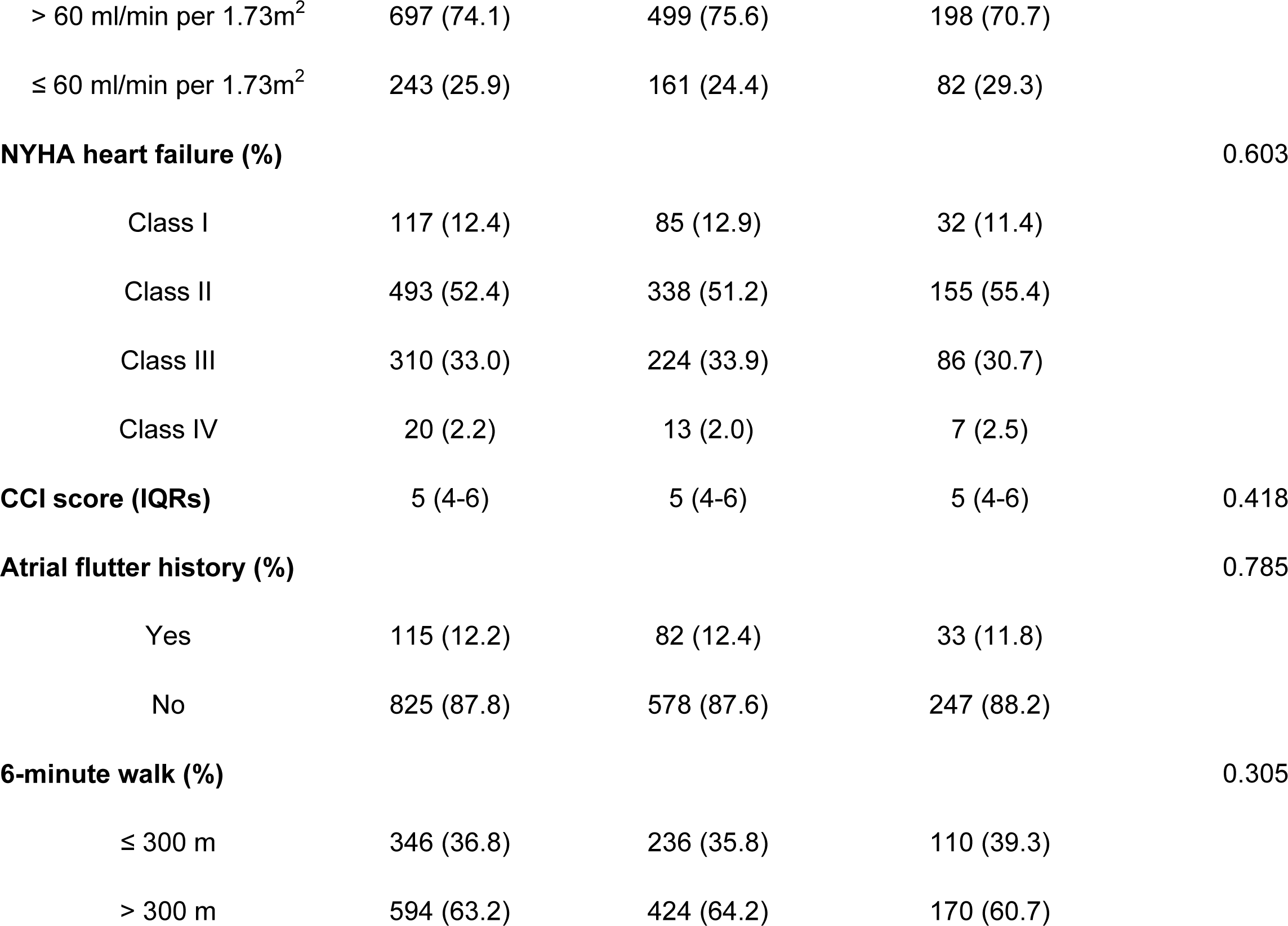
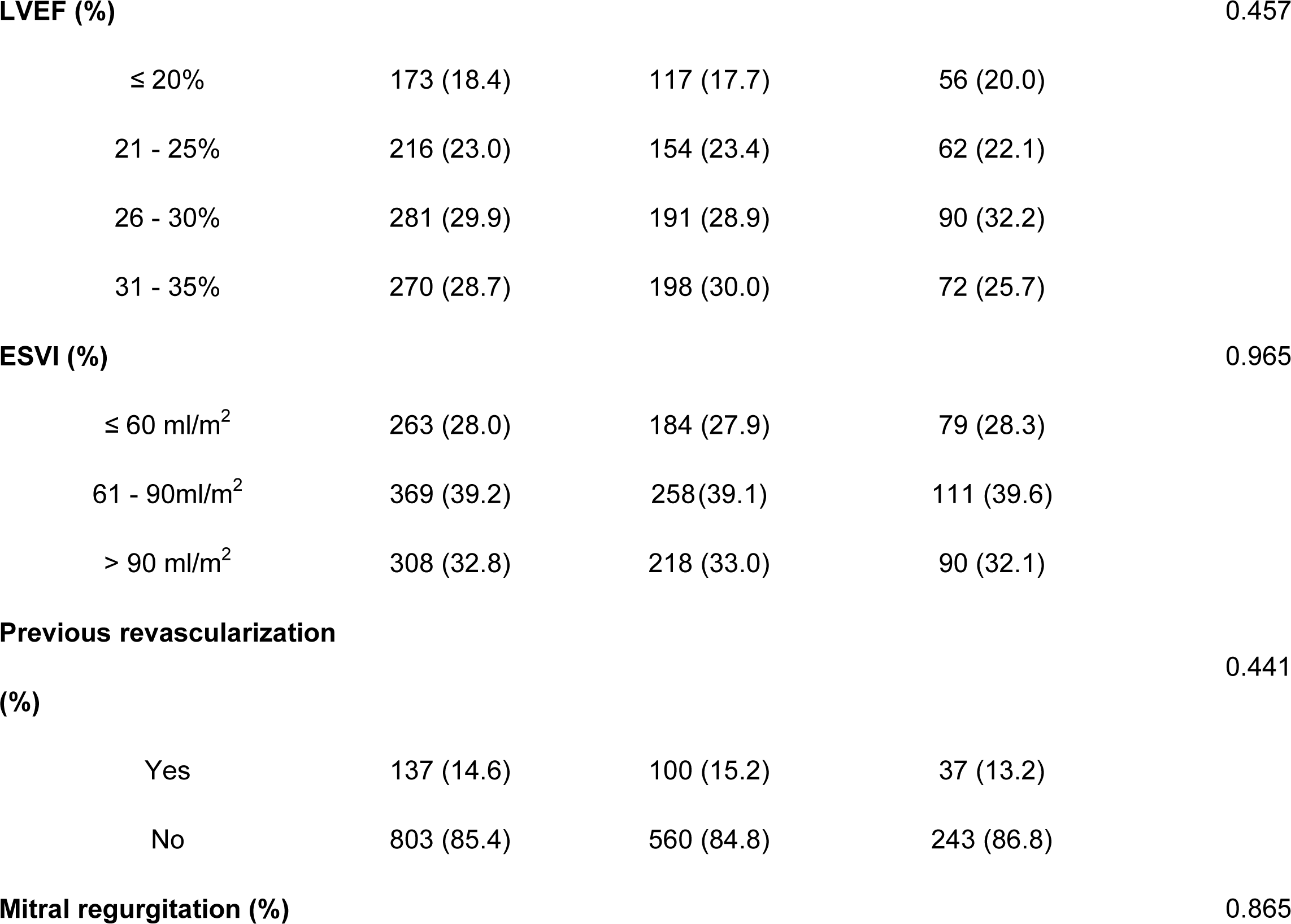
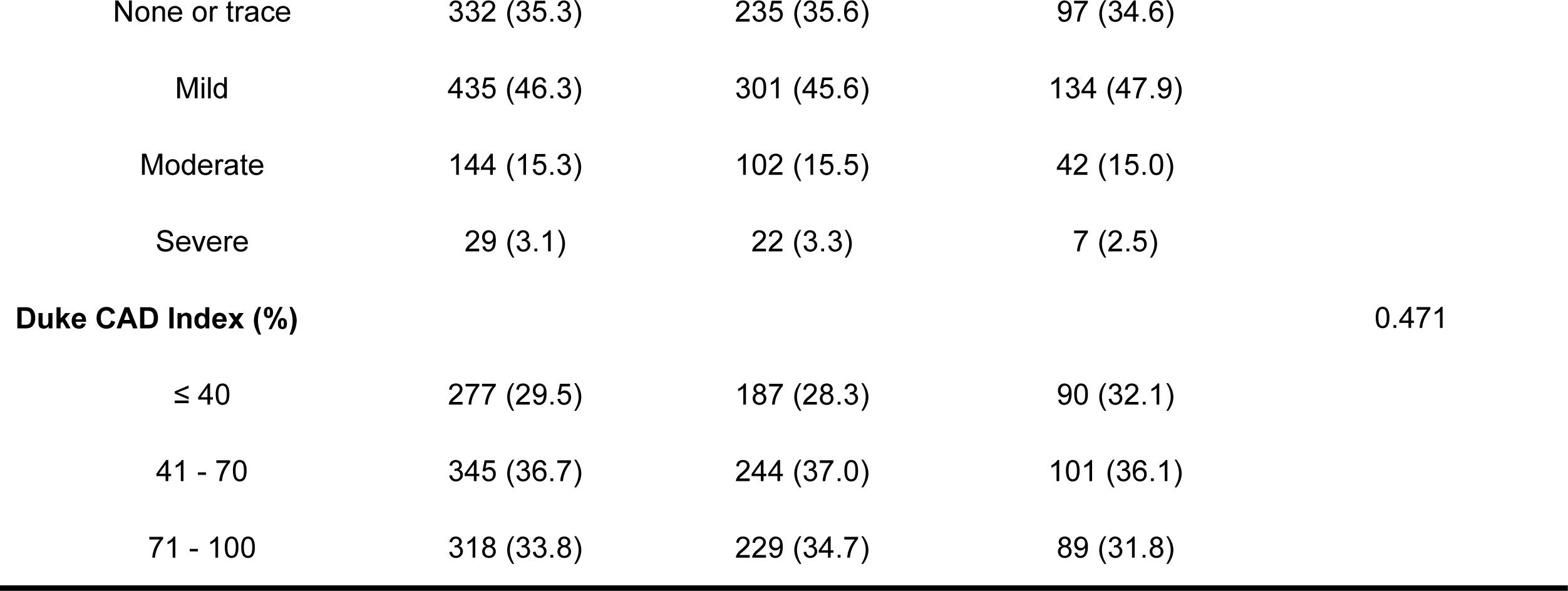
Baseline characteristics for entire cohort, training and validation groups. eGFR = estimated glomerular filtration rate; NYHA = New York Heart Association; CCI = Charlson co-morbidity index; LVEF = left ventricular ejection fraction; ESVI = end-systolic volume index; CAD = coronary artery disease.

| **Variables** | **Overall**<br>**(n = 940)** | **Training group**<br>**(n = 660)** | **Validation group**<br>**(n = 280)** | **p value** |
| --- | --- | --- | --- | --- |
| **Age (%)** |  |  |  | 0.564 |
| ≤ 50 | 139 (14.8) | 103 (15.6) | 36 (12.9) |  |
| 51 - 60 | 384 (40.9) | 273 (41.4) | 111 (39.6) |  |
| 61 - 70 | 284 (30.2) | 193 (29.2) | 91 (32.5) |  |
| ≥ 71 | 133 (14.1) | 91 (13.8) | 42 (15.0) |  |
| **Gender (%)** |  |  |  | 0.603 |
| Male | 828 (88.1) | 579 (87.3) | 249 (88.9) |  |
| Female | 112 (11.9) | 81 (12.3) | 31 (11.1) |  |
| **Ethnicity (%)** |  |  |  | 0.610 |
| White | 639 (68.0) | 452 (68.5) | 187 (66.8) |  |
| Other | 301 (32.0) | 208 (31.5) | 93 (33.3) |  |
| **eGFR (%)** |  |  |  | 0.117 |
| > 60 ml/min per 1.73m<sup>2</sup> | 697 (74.1) | 499 (75.6) | 198 (70.7) |  |
| ≤ 60 ml/min per 1.73m<sup>2</sup> | 243 (25.9) | 161 (24.4) | 82 (29.3) |  |
| **NYHA heart failure (%)** |  |  |  | 0.603 |
| Class I | 117 (12.4) | 85 (12.9) | 32 (11.4) |  |
| Class II | 493 (52.4) | 338 (51.2) | 155 (55.4) |  |
| Class III | 310 (33.0) | 224 (33.9) | 86 (30.7) |  |
| Class IV | 20 (2.2) | 13 (2.0) | 7 (2.5) |  |
| **CCI score (IQRs)** | 5 (4-6) | 5 (4-6) | 5 (4-6) | 0.418 |
| **Atrial flutter history (%)** |  |  |  | 0.785 |
| Yes | 115 (12.2) | 82 (12.4) | 33 (11.8) |  |
| No | 825 (87.8) | 578 (87.6) | 247 (88.2) |  |
| **6-minute walk (%)** |  |  |  | 0.305 |
| ≤ 300 m | 346 (36.8) | 236 (35.8) | 110 (39.3) |  |
| > 300 m | 594 (63.2) | 424 (64.2) | 170 (60.7) |  |
| **LVEF (%)** |  |  |  | 0.457 |
| ≤ 20% | 173 (18.4) | 117 (17.7) | 56 (20.0) |  |
| 21 - 25% | 216 (23.0) | 154 (23.4) | 62 (22.1) |  |
| 26 - 30% | 281 (29.9) | 191 (28.9) | 90 (32.2) |  |
| 31 - 35% | 270 (28.7) | 198 (30.0) | 72 (25.7) |  |
| **ESVI (%)** |  |  |  | 0.965 |
| ≤ 60 ml/m<sup>2</sup> | 263 (28.0) | 184 (27.9) | 79 (28.3) |  |
| 61 - 90ml/m<sup>2</sup> | 369 (39.2) | 258 (39.1) | 111 (39.6) |  |
| > 90 ml/m<sup>2</sup> | 308 (32.8) | 218 (33.0) | 90 (32.1) |  |
| **Previous revascularization** |  |  |  |  |
| **(%)** |  |  |  | 0.441 |
| Yes | 137 (14.6) | 100 (15.2) | 37 (13.2) |  |
| No | 803 (85.4) | 560 (84.8) | 243 (86.8) |  |
| **Mitral regurgitation (%)** |  |  |  | 0.865 |
| None or trace | 332 (35.3) | 235 (35.6) | 97 (34.6) |  |
| Mild | 435 (46.3) | 301 (45.6) | 134 (47.9) |  |
| Moderate | 144 (15.3) | 102 (15.5) | 42 (15.0) |  |
| Severe | 29 (3.1) | 22 (3.3) | 7 (2.5) |  |
| **Duke CAD Index (%)** |  |  |  | 0.471 |
| ≤ 40 | 277 (29.5) | 187 (28.3) | 90 (32.1) |  |
| 41 - 70 | 345 (36.7) | 244 (37.0) | 101 (36.1) |  |
| 71 - 100 | 318 (33.8) | 229 (34.7) | 89 (31.8) |  |

### 3.2. Identification of the independent predictors

Univariate and multivariate Cox proportional hazards models were performed to identify the independent predictors for OS in the training group. In the multivariate analysis, seven variables, including treatment type (P = 0.032), gender (P < 0.001), eGFR (P = 0.018), CCI score (P = 0.002), 6-minute walk (P = 0.040), ESVI (P = 0.006) and MR class (P < 0.001) were identified as the independent predictors for OS (Table 2).

**Table 2.**
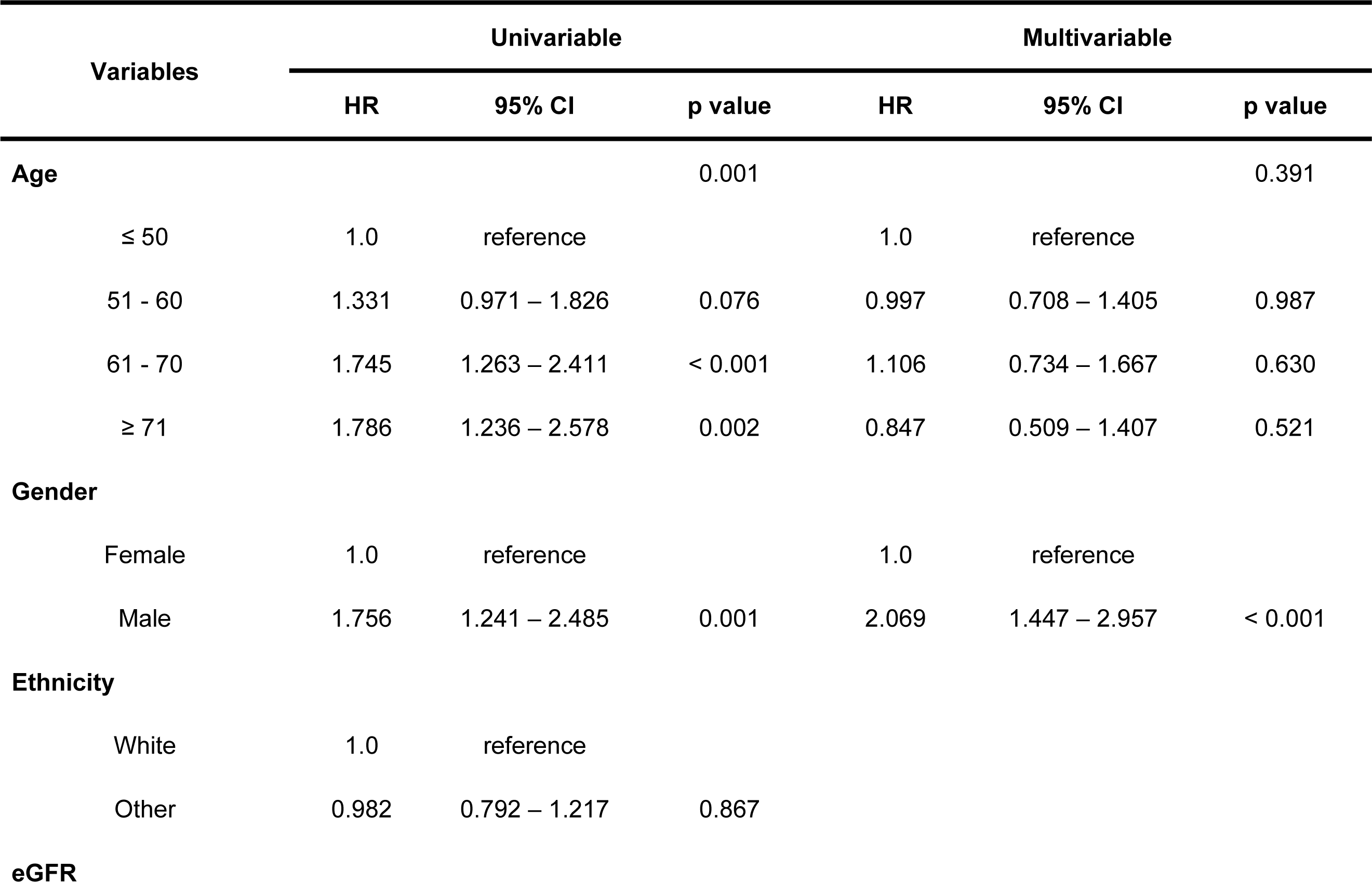
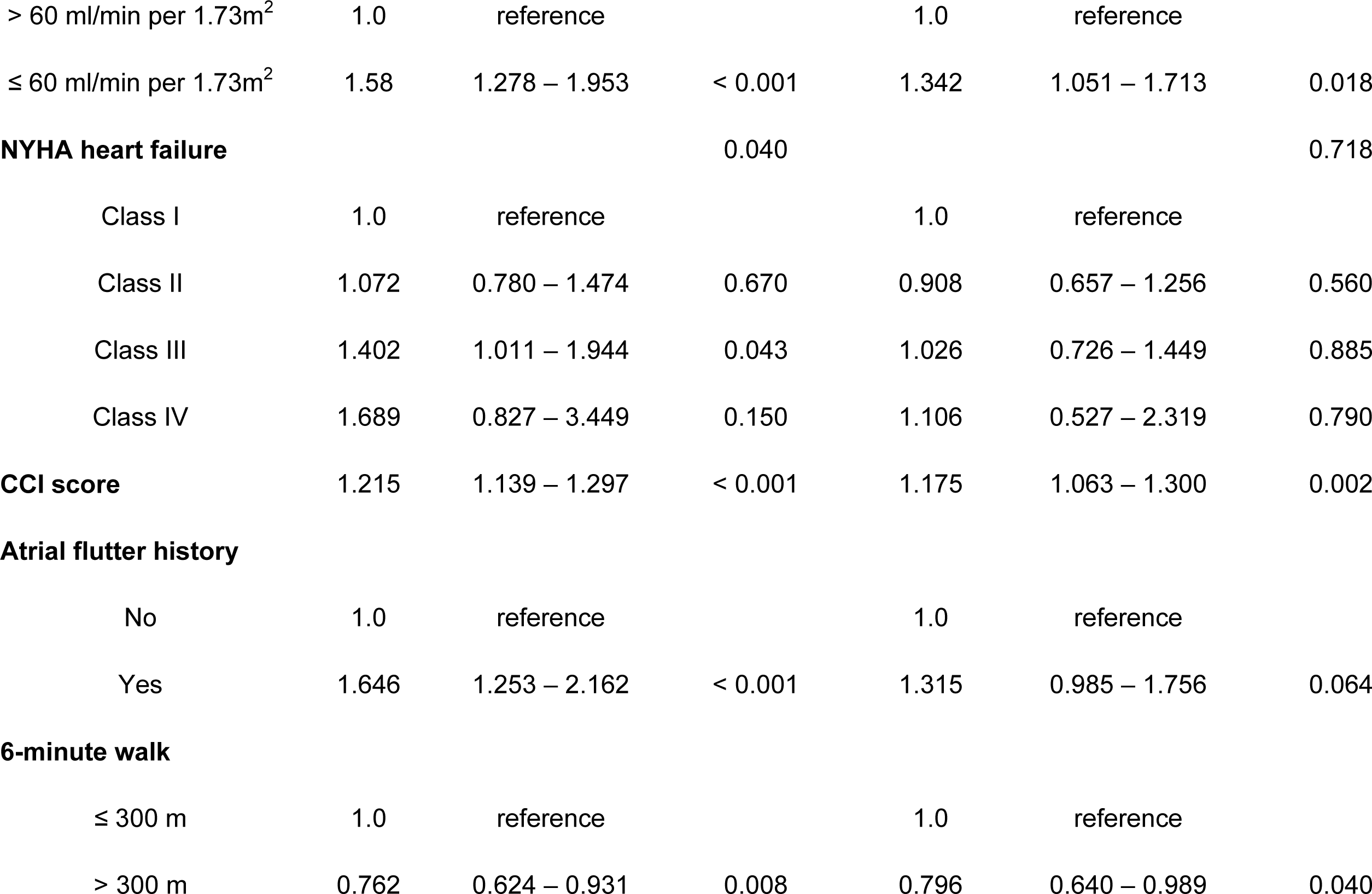
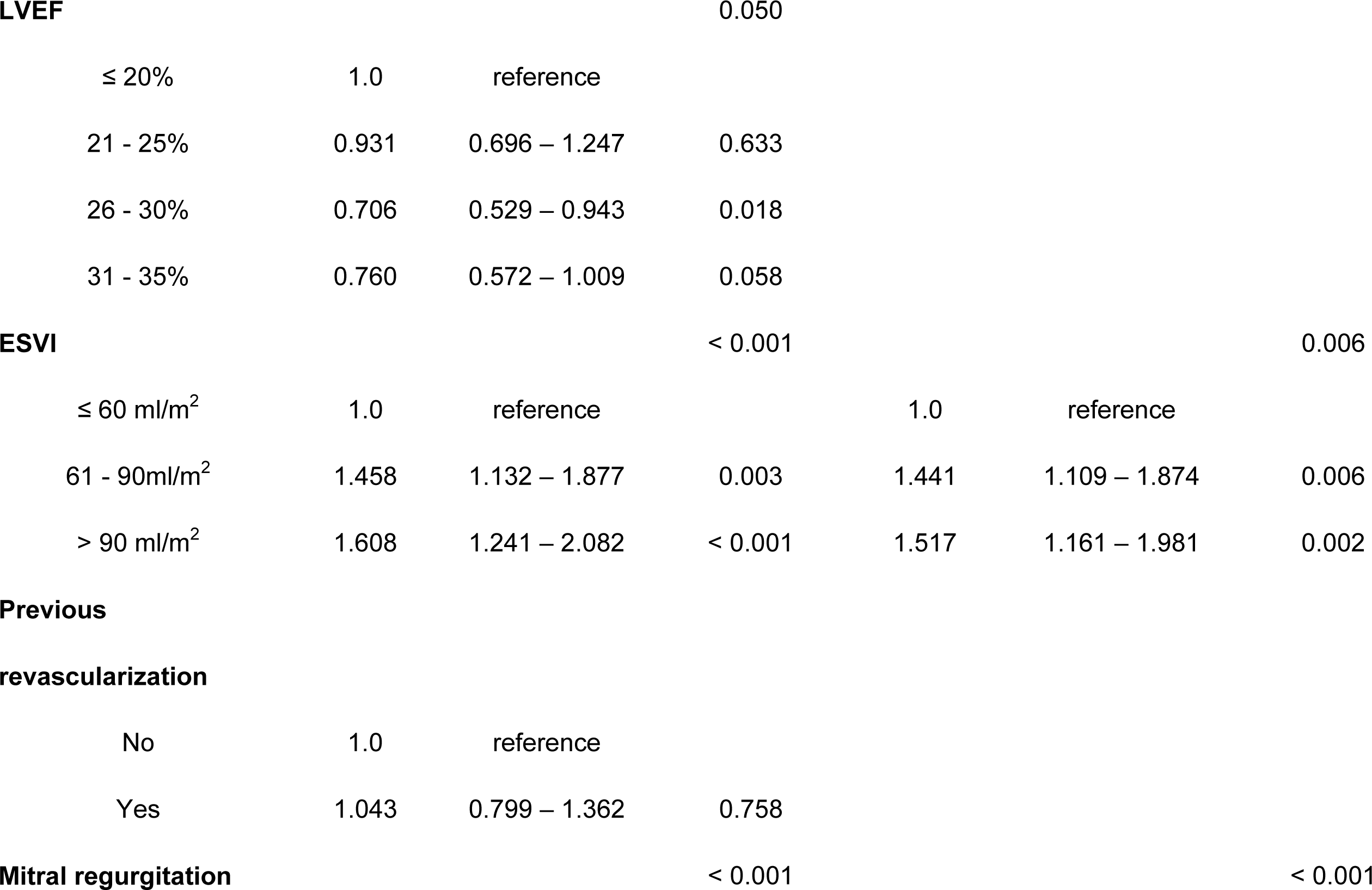
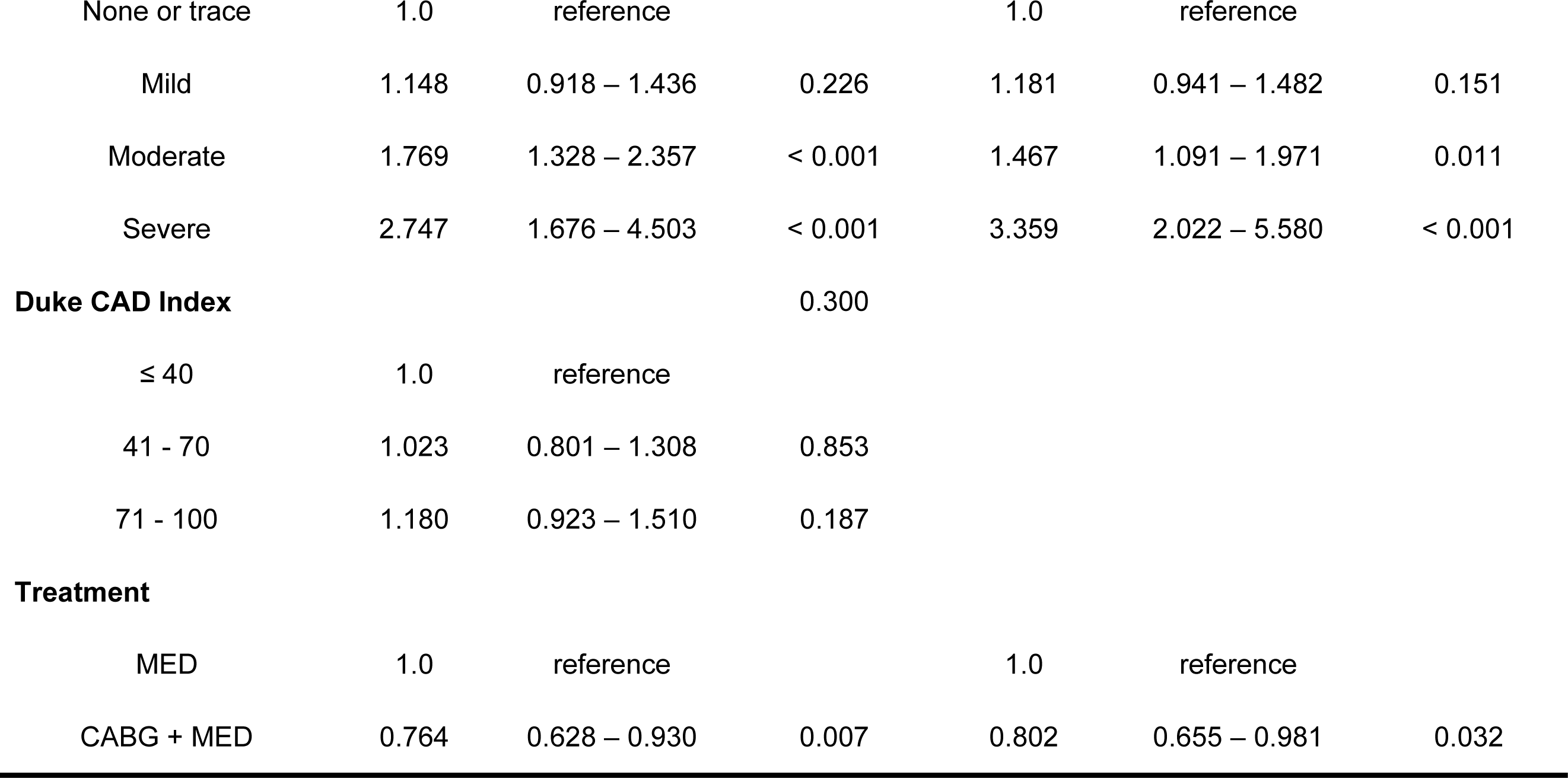
Univariate and multivariate Cox proportional hazards models of OS for patients with ischemic cardiomyopathy and heart failure in the training group. HR = hazards ratio; CI = confidence interval; eGFR = estimated glomerular filtration rate; NYHA = New York Heart Association; CCI = Charlson co-morbidity index; LVEF = left ventricular ejection fraction; ESVI = end-systolic volume index; CAD = coronary artery disease; CABG = coronary artery bypass grafting; MED = medical.

### 3.3. Nomogram establishment and verification

The 1-year, 3-year, 5-year, and ten-year predictive nomogram of OS for patients with ICM and heart failure were established after incorporating the seven independent predictors in the multivariate Cox proportional hazards model. (Figure. 2) Using bootstraps with 1000 resamples, the C-indices of the nomogram were 0.641 (95% CI: 0.627-0.655) and 0.649 (95% CI: 0.627-0.671) for training and validation groups, respectively. The predicted and actual risks of 1-year, 3-year, 5-year, and ten-year OS were consistent as shown in the calibration curves (Figure. 3). The values of area under curve (AUC) were 0.634, 0.616, 0.630 and 0.638 for 1-year, 3-year, 5-year and ten-year ROC curves in the training group, respectively (Figure. 4). DCA curves identified the 1-year, 3-year, 5-year and ten-year net benefits during the clinical application of the nomogram in decision-making for patients with ICM and heart failure in training and validation groups (Figure. 5). Of note, a tangible elevated trend of net benefit was shown with the extension of predicted nodal time in DCA curves. The prognostic index of 1-year OS for each patient calculated by the independent predictors and corresponding hazards ratio, and the cutoff values of prognostic indices were 6.698 and 6.833 for training and validation groups, respectively. Based on the respective cutoff values, the two groups were stratified into high-risk and low-risk groups, respectively. The Kaplan-Meier survival analysis identified better OS among patients with low-risk group compared to those with high-risk group in training and validation groups (both P < 0.0001), confirming the excellent discriminatory power of the nomogram (Figure. 6).

**Figure 2.**
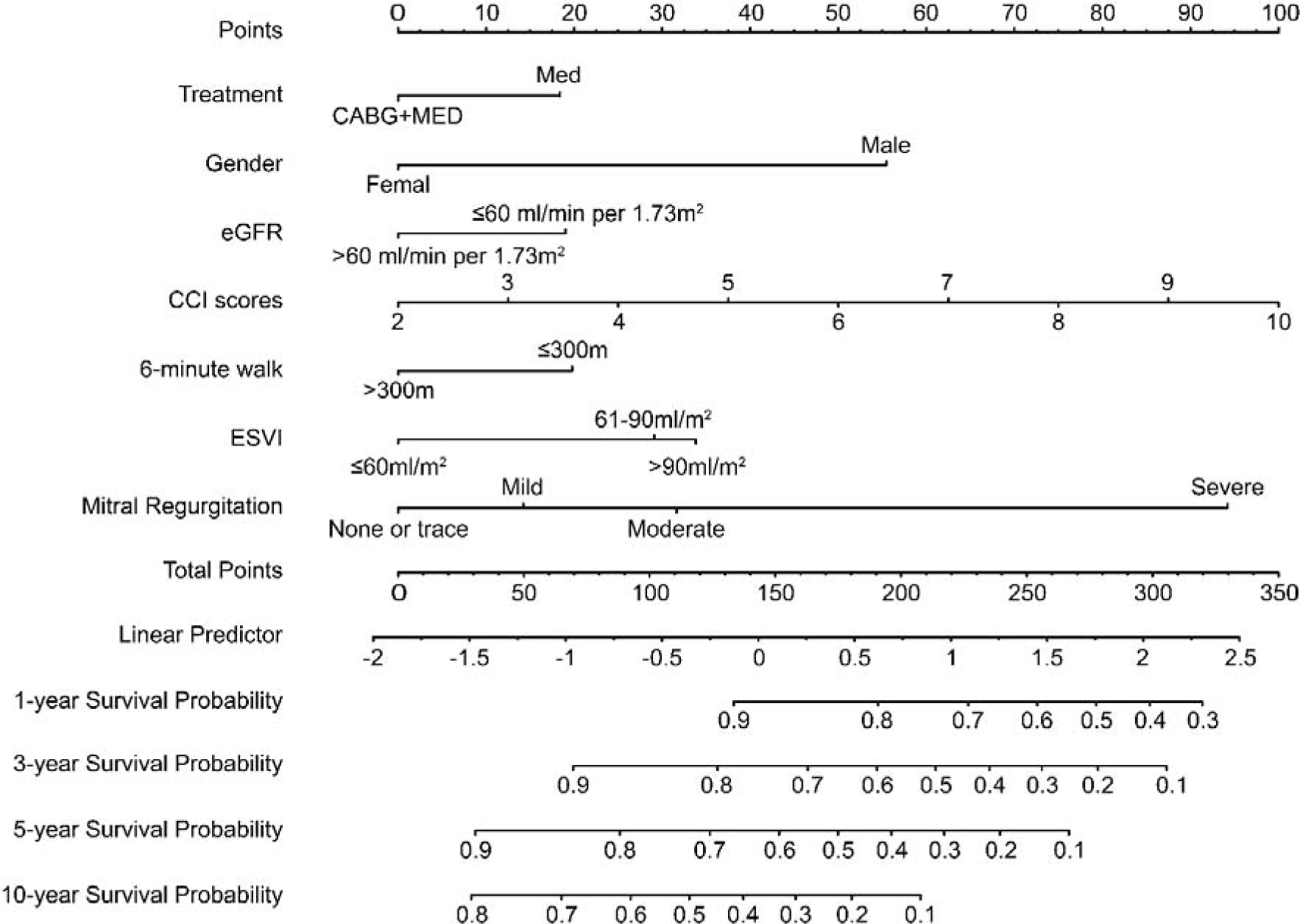
Nomogram for 1-year, 3-year, 5-year, and ten-year prediction of overall survival for patients with ischemic cardiomyopathy and heart failure. CABG = coronary artery bypass grafting; MED = medical; eGFR = estimated glomerular filtration rate; CCI = Charlson co-morbidity index; ESVI = end-systolic volume index.

**Figure 3.**
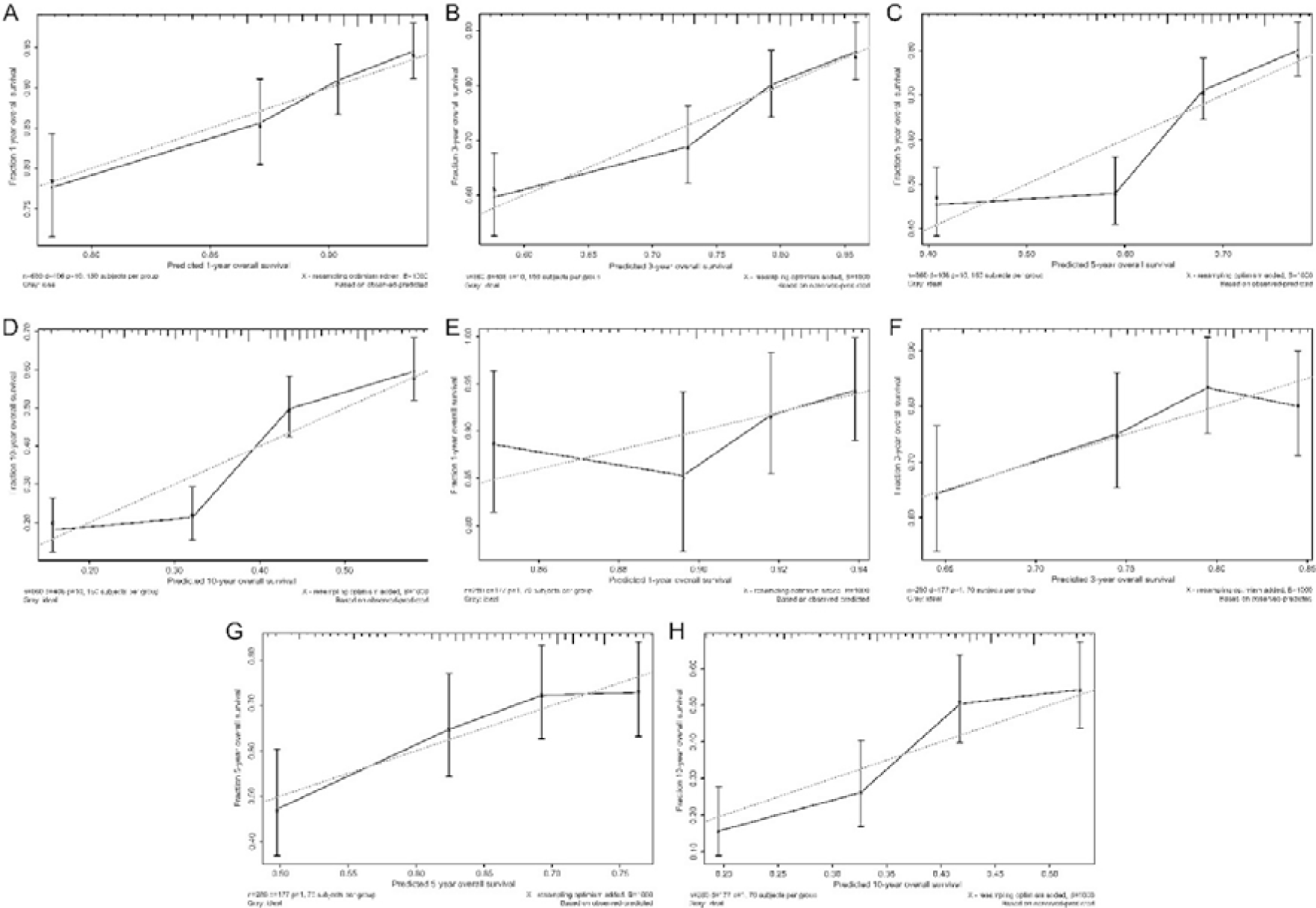
Calibration curves of 1-year, 3-year, 5-year, and ten-year overall survival (OS) for patients with ischemic cardiomyopathy and heart failure. A: 1-year OS in training group; B: 3-year OS in the training group; C: 5-year OS in the training group; D: ten-year OS in the training group; E: 1-year OS in the validation group; F: 3-year OS in the validation group; G: 5-year OS in the validation group; H: ten-year OS in the validation group.

**Figure 4.**
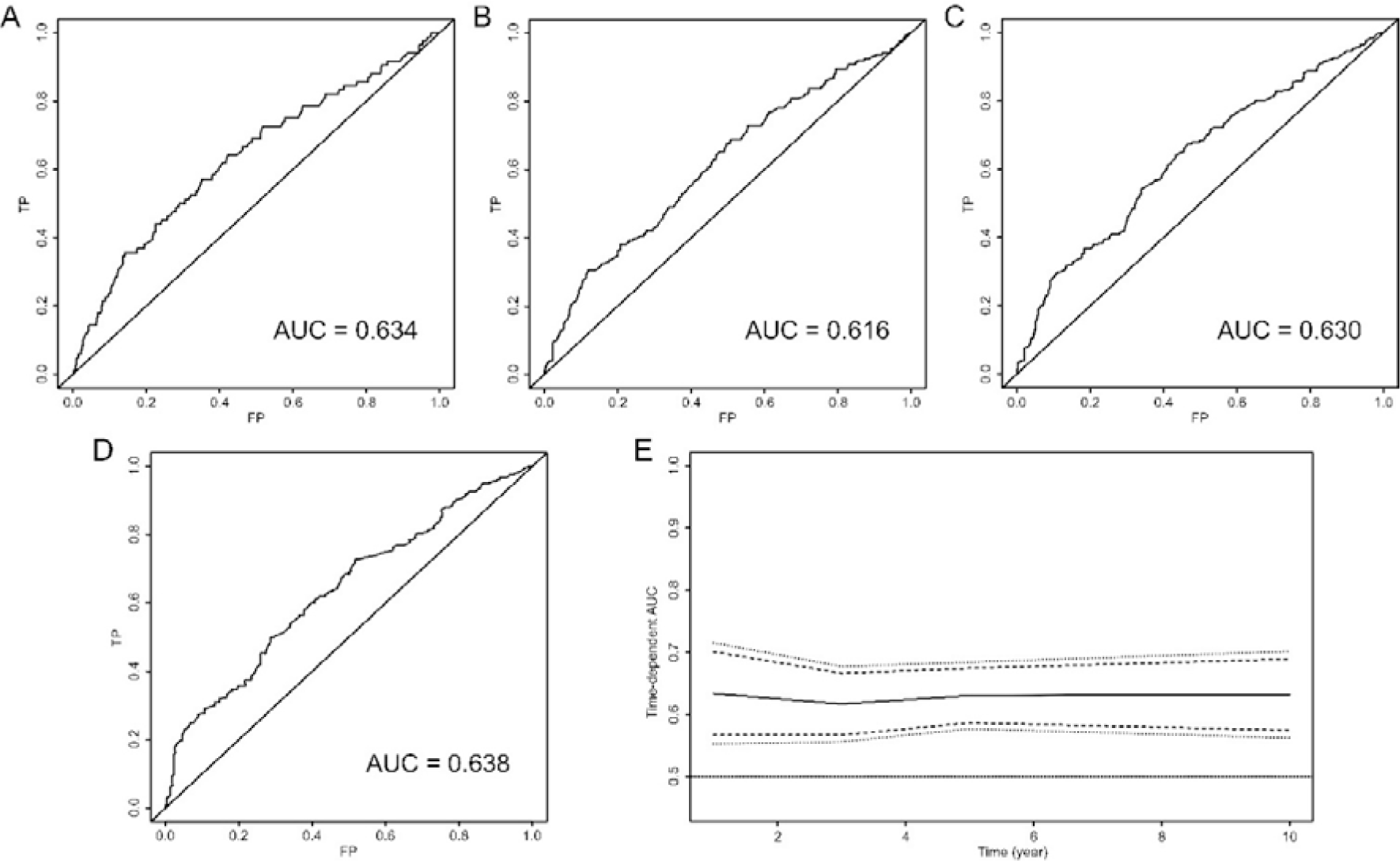
Time-dependent receiver operating characteristic curves of the overall survival (OS) in the training group for patients with ischemic cardiomyopathy and heart failure. A: 1-year OS; B: 3-year OS; C: 5-year OS; D: ten-year OS; E: time-dependent area under curve (AUC).

**Figure 5.**
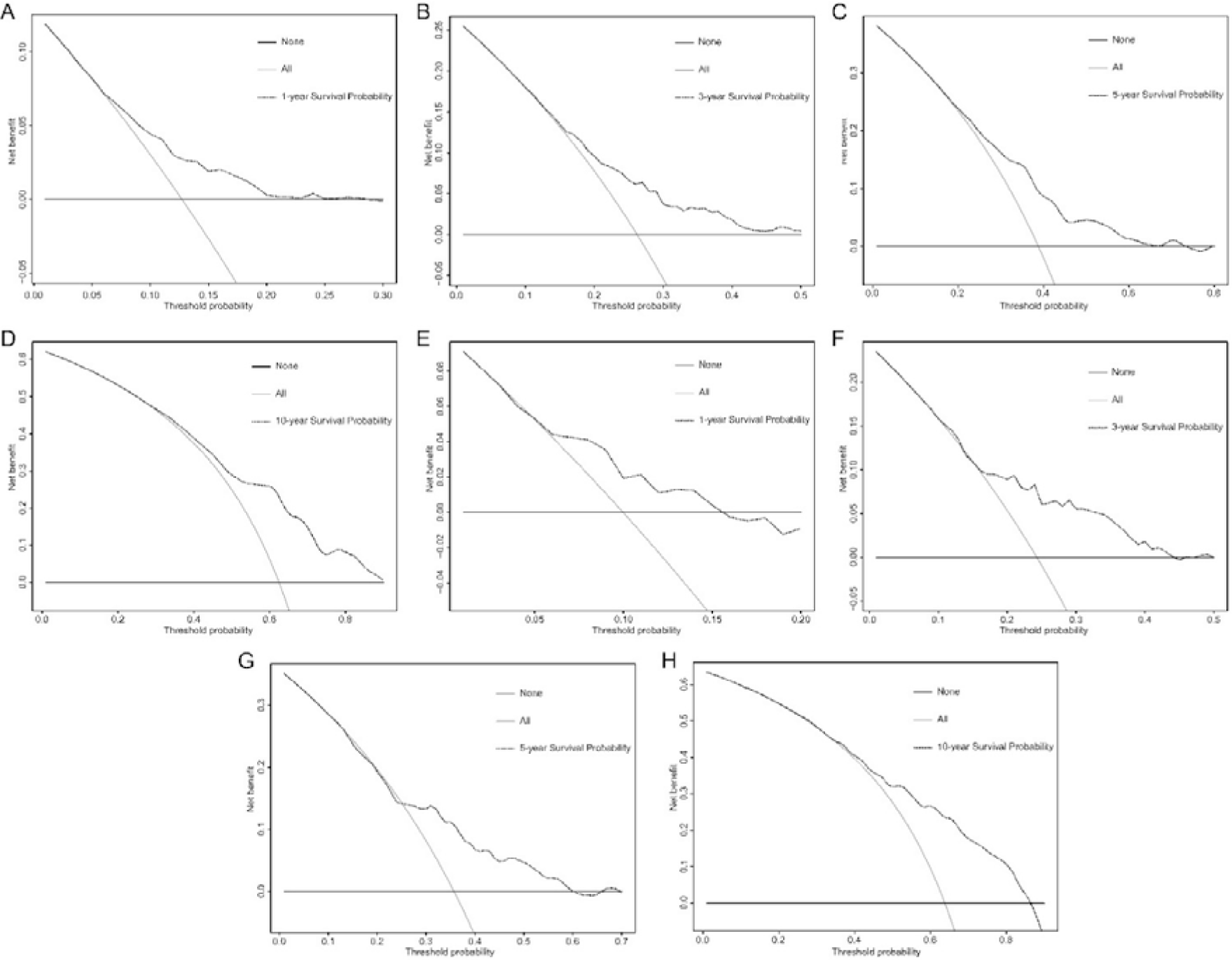
Decision Curve Analysis of the nomogram in training group (A: 1-year OS, B: 3-year OS, C: 5-year OS, D: ten-year OS) and verification group (E: 1-year OS, F: 3-year OS, G: 5-year OS, H: ten-year OS). The horizontal black solid lines assumed all cases were alive, and the oblique gray solid lines assumed all cases were dead. The black dash lines represented net benefit of the model during the application in clinical decision-making. OS = overall survival.

**Figure 6.**
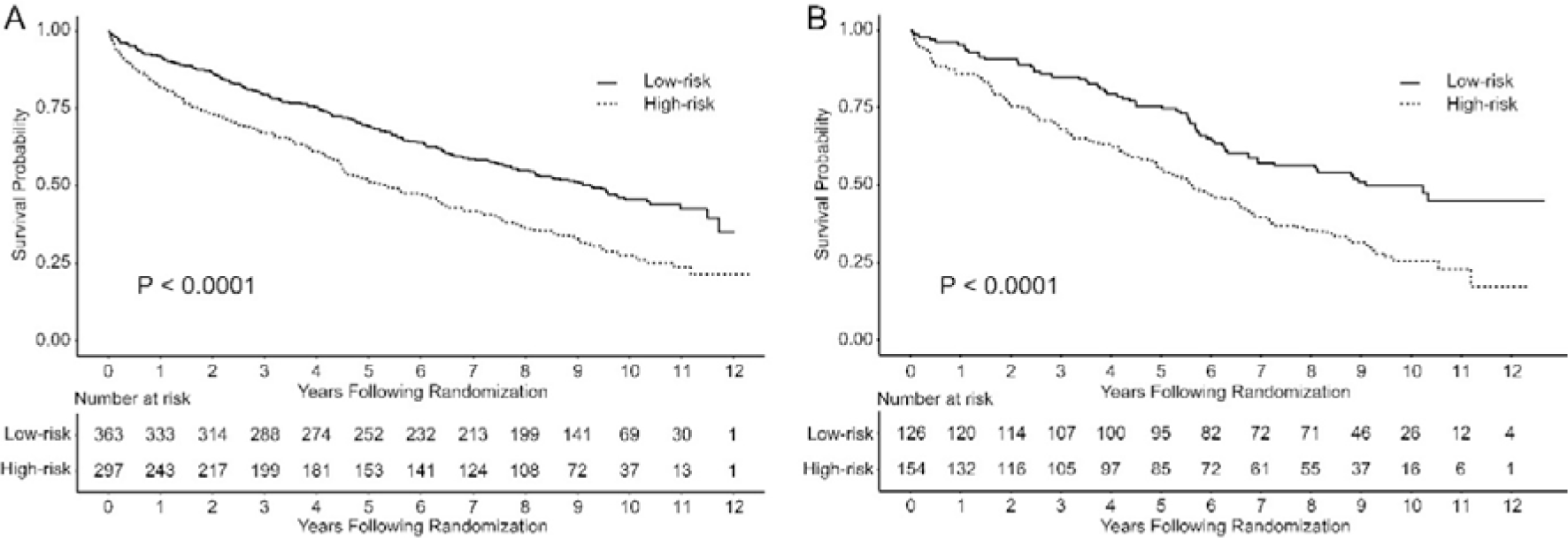
Kaplan-Meier survival analysis of overall survival among patients with low-risk group and patients with high-risk group in training and validation groups. A: training group; B: validation group.

## 4. Discussion

The cardinal principle of management of patients with ICM and heart failure is to improve the health status-related quality of life and prolong lifespan. However, in light of the dismal prognosis and scant randomized trails, divergent viewpoints have evolved among practitioners when treating this specific cohort. From this perspective, a rigorous selection process with the assistance of a specialized model covering universal and homogeneous variables among these patients to quantify the survival probability and optimize the prognostic discrimination should be implemented during counselling for clinical decision-making.

Several strengths of current nomogram are noteworthy. First, the current nomogram may inform practice in an area where, up to now, there has been no specialized model to predict overall survival for patients with ICM and heart failure. Second, the enrolled variables including the evaluation of cardiac function, physical status, comorbidities, demographics, and treatment type are multifaceted and easily accessible, and on this premise, the current nomogram may provide ease of use in the clinical setting. Third, considering the mutual causality between renal insufficiency and heart failure, according to the recommendation of the Kidney Disease: Improving Global Outcomes (KDIGO) organization, the estimated glomerular filtration rate calculated by CKD-EPI equation was endorsed to precisely evaluate the renal function instead of creatinine or creatinine clearance rate.^18^ Fourth, it was reported more than 70% of patients enrolled in the STICH trail had a severe burden of medical co-morbidities at baseline, resulting in a ten-year mortality of approximately 70% in that cohort.^19^ Therefore, in the current study, the available medical co-morbid conditions among these fragile patients were numerically quantified by the CCI in an effort to provide the linear relation of risk between CCI score and OS. Fifth, in light of the tangible elevated trend of net benefit shown with the extension of predicted nodal time in DCA curves, the current nomogram can be used as a supplement to the STS and EuroSCORE II risk models for predicting long-term survival. Taken as a whole, the aforementioned strengths endowed practicality and generalizability to current nomogram in clinical decision making with the engagement of clinicians and patients. The twice risk of mortality for male compared to female deserves attention in current study. Our result was in agreement with the finding of Pina et.al, who identified a significant diminished all-cause mortality of 33% lower among female compared to male based on STICH trail.^20^ The extended follow-up of the MAIN-COMPARE (Ten-Year Outcomes of Stents Versus Coronary-Artery Bypass Grafting for left Main Coronary-Artery Bypass Grafting for Left Main Coronary Artery Disease) trail also revealed lower all-cause mortality and serious composite outcomes at 10 years in female compared with male, and this difference was mainly driven by the higher event rate in male during the late period between 5 and 10 years.^21^ On the contrary, multiple lines of evidence have documented the poorer prognosis of female compared to male due to various pathologic resultant such as medical co-morbid conditions, coronary microvascular dysfunction and diastolic filling pattern.^22–24^. However, the 2021 ACC/AHA/SCAI guideline for coronary artery revascularization highlighted that treatment decisions for patients with CAD requiring revascularization should be based on clinical indications, regardless of gender, race, or ethnicity in an effort to mitigate disparities of care.^2^ Yoon et.al reported that for elderly patients with ICM who had a LVEF < 40% and left ventricular end diastolic dimension > 55mm, different genders represented distinctly predominant series of risk factors of mortality.^25^ On this premise, the aforementioned perspectives were not inevitable in contrast to our result, and this lack of consistency should be expounded in the setting of heterogeneous confounders with respect to study cohorts, genetics, neuroendocrine activation, anatomic complexity, medications prescribed and their response to the disease process, and weights of variables.^26^ Actually, continuing debate still urges additional attempts to address a consensus surrounding the interaction between gender and long-term survival in patients with CAD.

Several important variables with regard to the evaluation of cardiac function were failed to contribute to the construction of the current nomogram after adjusting for baseline characteristics. Undoubtedly, the hierarchical thresholds of LVEF can be used as a perfect tool to identify different subsets of patients with CAD, who have various degrees of damaged left ventricular segmental systolic function. This notwithstanding, it may not portend that they can invariably provide sufficient efficiency to predict survival for some specific cohorts. The current study was in agreement with several contemporary studies that had revealed inability of LVEF to shed light on short-term and long-term survival in patients with severe heart failure.^27–30^ Withal, the underestimated LVEF caused by MR that accounted for 65% patients in the current cohort might also contribute to the negative statistical power. Indeed, in addition to LVEF, a serious of cardiac substrate measured by echocardiography that are independent of LVEF, such as E/A ratio, severity of MR, sphericity index, ESVI, end-diastolic volume index, pulmonary artery pressure and mechanical dyssynchrony, may typically present various degrees of adverse changes accompanied by progressive left ventricular dilatation and adverse remodeling, and have been identified as the independent risk factors for mortality.^30^ However, due to the large number of missing values in the STICH data, some of aforementioned variables were not analyzed, thus adding residual confounding to current study.

Another indolent variable was NYHA heart failure classification which conferred a graded risk of mortality in the univariate Cox model but was blunted in multivariate analysis. This unexpected result ran counter to the prevailing wisdom of accumulated dogmas suggesting NYHA heart failure classification as a foundational criterion for risk stratification. However, the variable subjectivity and poor reproducibility have raised concerns about the accuracy of NYHA heart failure classification in evaluating prognosis. The findings of Caraballo et al. who conducted secondary analyses of four multicenter National Institutes of Health-funded heart failure clinical trials demonstrated poor discrimination across the spectrum of functional impairment and unreliable prediction of survival, based on NYHA heart failure classification.^31^ Greene et al. compared the NYHA heart failure classification and the Kansas City Cardiomyopathy Questionnaire Overall Summary Score (KCCQ-OS) with respect to clinically meaningful changes in health status over time, and found that changes in NYHA heart failure classification failed to translated into a benefit of clinical outcomes with respect to all-cause mortality, heart failure hospitalization, and these two composition.^32^ Obviously, the STICH trail inevitably suffered the aforementioned limitations from the fact that patients were recruited from 99 medical centers in 22 countries.

The current study can provide actionable evidence for contemporary practice in the following aspects. First, our results enhanced the consensus that CABG in conjunction with guideline-directed medical therapies could be the preferred recommendation for patients with ICM and heart failure by reducing 20% risk of all-cause mortality compared to medical therapy alone. Second, on the basis of concern about the high weight of comorbidities in current nomogram, a comprehensive multidisciplinary therapeutic is warranted and may potentially improve the survival time. Third, given the reversibility of ischemic MR, strategies to reduce the severity of MR are urgent in an effort to reduce the mortality, whether or not CABG or medical therapy alone is selected. Fourth, the current nomogram in conjunction with the STS and EuroSCORE II risk models may serve as a perfect tool in clinical decision making with the engagement of clinicians and patients.

Our study cannot avoid certain inevitable limitations due to the post-hoc nature and lack of external validation. Due to the large number of missing values, several vital variables were not collected, and selection biases added insufficiently corrected residual confounding to current study. The small size of subgroups of MR and NYHA heart failure classification can be translated into heterogeneity, compromising the reliability of the nomogram. The fact that several medical co-morbidities were not recorded in the STICH trail caused the underestimated CCI scores. Based on the STICH protocol, the generalizability of current nomogram might not be suitable for patients who have the conditions among exclusion criteria. Therefore, our results should be interpreted with caution and external validation is warranted.

## 5. Conclusion

The current study developed a specified nomogram containing seven multifaceted and easily accessible variables to predict the OS for patients with ICM and heart failure, and provided information about multidisciplinary therapeutic that may prolong the survival time. This nomogram in conjunction with the STS and EuroSCORE II risk models may serve as a perfect tool in clinical decision making with the engagement of clinicians and patients.

## Data Availability

The data can be accessed via the Biologic Specimen and Data Repository Information Coordinating Center after the approval of the National Heart, Lung, and Blood Institute.

https://biolincc.nhlbi.nih.gov

## Nonstandard Acronyms and Abbreviations

OS: overall survival
ICM: ischemic cardiomyopathy
CABG: coronary artery bypass grafting
PCI: percutaneous coronary intervention
LVEF: left ventricular ejection fraction
CAD: coronary artery disease
NYHA: New York Heart Association
MR: mitral regurgitation
ESVI: end-systolic volume index
eGFR: estimated glomerular filtration rate
CKD-EPI: Chronic Kidney Disease Epidemiology Collaboration
CCI: Charlson co-morbidity index
MED: medical
IQR: interquartile range
ROC: receiver operating characteristic
DCA: Decision Curve Analysis
HR: hazards ratio
CI: confidence interval
AUC: area under curve

## Acknowledgments

Conception and design: Pengju Guo and Chang He. Acquisition of data: Pengju Guo. Analysis and interpretation of data: Pengju Guo and Chang He; Manuscript writing: Pengju Guo. Revision: Junlei LI, Youxu Jiang, Feng Wang, Jiaxiang Wang, Bin Lin, and Deguang Feng. All the authors listed have approved the manuscript that is enclosed and agree with its content.

